# Immunity after COVID-19: protection or sensitization?

**DOI:** 10.1101/2020.05.21.20108860

**Authors:** Antoine Danchin, Gabriel Turinici

**Affiliations:** Kodikos Labs, Institut Cochin, 75014, Paris, FRANCE; CEREMADE, Université Paris Dauphine - PSL, Paris 75016, FRANCE

**Keywords:** COVID-19, antibody-dependent enhancement, SEIR model, vaccine, dengue fever, asymptomatic, **MSC codes:** 92C60, 92D30, 34C60

## Abstract

Motivated by historical and present clinical observations, we discuss the possible unfavorable evolution of the immunity (similar to documented antibody-dependent enhancement scenarios) after a first infection with COVID-19. More precisely we ask the question of how the epidemic outcomes are affected if the initial infection does not provide immunity but rather sensitization to future challenges. We first provide background comparison with the 2003 SARS epidemic. Then we use a compartmental epidemic model structured by immunity level that we fit to available data; using several scenarios of the fragilization dynamics, we derive quantitative insights into the additional expected numbers of severe cases and deaths.

## 1 Introduction

The present outbreak of COVID-19 and the unprecedented response from the global human population, whether at an individual level or collectively at a political level, integrate a myriad of factors. Making out the most relevant lessons from the course taken by the disease will take time. In particular many perplexing questions have not yet found adequate answers, that would be crucial for the design of optimal treatment policies and healthcare strategies, both at the individual and societal level. Among these questions, prominent ones are: Why are the mortality patterns so skewed toward higher ages? What is the common denominator of the observed comorbidities that affect in a non-negligible way the outcome of the disease? What is the size of the asymptomatic cohorts? Are asymptomatic (or pauci-symptomatic) cases a barrier or an accelerator of the disease propagation? etc. To these salient questions, which deal with the present situation, we should add questions relative to future outbreaks, their severity and characteristics. This is particularly important as health authorities have, for obvious reasons, focused on the design of vaccines, despite the fact that vaccines are sometimes extremely difficult to obtain [19]. In particular while a response to previous infections is usually protective it can sometimes be deleterious and result in severe outcomes (see e.g. [17]). Answering many of these questions requires deep understanding of the immune response of the host, whether innate or acquired, at the cellular or humoral level. Recent research pointed out that it is critical to investigate previous history of interaction between host and infective virus strains (see [2] for SARS-CoV-2 and [13] for the Zika virus). In this context, the question of whether previous infection with coronaviruses is beneficial or detrimental to the immunity of the host is a matter of active debate, see for instance [3, 23, 24].

Here, we focus on the so-called *antibody-dependent enhancement* (ADE) mechanism (related to the more colorful name of *cytokine storm)*. ADE corresponds to a situation where antibodies that normally alleviate the consequences of a viral infection end up doing the opposite: they fail to control the virus’ pathogenicity by failing to be neutralizing (i.e., the antibodies are not able to kill the virus), or even enhance its virulence either by facilitating its entry into the cell (thus enhancing the viral reproduction potential), or by triggering an extensive and misadapted reaction, thereby causing damage to the host organs through hyper-inflammation (cytokine storm).

Several documented examples are known that give rise to ADE: in SARS (see [4]), dengue (see [9, 12]), HIV-1 [30] to cite but a few. This is even already documented in the case of COVID-19 as the similarity with dengue fever has resulted in false-positive identification when using diagnostic tests based on serology (see for example in Singapore [31]). Furthermore, recent genomic investigations [26] show that reinfection can indeed occur (as opposed to viral shedding from the primary infection). Let us illustrate the first two situations more in detail: for dengue fever it is established that a previous infection with a virus belonging to a particular serotype family can cause adverse reaction upon re-infection with a virus from another serotype (see [9]). This unwanted outcome was demonstrated in a study that monitored two dengue fever outbreaks: a first one in Cuba in 1977 [5], followed by a second one twenty years later (1997) [10]. Being infected during the first epidemic was proven to negatively influence the outcome of patients infected in the 1997 epidemic (with a dengue fever serotype that differed from that of 1997). To put this otherwise, the acquired immune response was fit for a given serotype but detrimental upon challenge with a different serotype. In the present context, the large time lapse between the two dates is worth emphasizing. Irrespective of whether or not the challenge is or is not related to a different serotype, we will call this situation a “Cuban hypothesis” and discuss it below.

A misadapted immune response is not limited to infection by the dengue virus (DENV). Another documented ADE case in animal models concerns the coronavirus family. M. Bolles and his collaborators (see [4]) tested a coronavirus vaccine on animal models and the results differed, depending on the age of the vaccinated animal: while the vaccine provided partial protection against both homologous and heterologous viruses for young animal models, the same vaccine performed poorly in aged animal models and was potentially pathogenic. This shows that a faulty immune response can depend on the general maturation of the immune system, with a significant age-dependent component.

At least four human coronaviruses [229E, NL63, OC43 and HKU1 [7]] are known to be endemic. They infect mainly the upper respiratory tract. To these we must add the now well-known strains SARS-CoV-1 (responsible for the 2003 epidemic), MERS-CoV and SARS-CoV-2. If a “Cuban hypothesis” is found to account for the persistent development of related diseases, the challenge by any one of these viruses or novel variants may reach human populations in a variety of forms. This dire anticipation prompted us to investigate the impact of such a scenario on the immune system status within the population at large. In this context, we ask here the following questions:

1. is the severity of the current individual outcomes influenced by previous coronavirus infections?;
2. what is the likely state of the immune system after a first SARS-CoV-2 infection?;
3. how can the present epidemic impact a future (homologous or heterologous, endemic or pandemic) coronavirus outbreak or any medical condition triggering the immune response induced by a primary infection?

## 2 Methods

### 2.1 The epidemic model of the impact of previous virus infection

The epidemic model we explored is the following: during the initial infection pathway, any patient starts in a “Susceptible” (notation “S”) class, can progress to the “Exposed” (notation “E”) class and then either to a “Severely infected” (label “*I_S_*”) or “Mild infected” class (including the asymptomatic and undetected individuals, denoted “*I_M_*”). The mild infected progress, after 1/γ*_M_* days to a “Mild Recovered” *”R_M_*” class while the “Infected” can either die (class “Deceased”, “D”) or recover after 1/γ*_I_* days and arrive in class “*R_S_*”. Note that, as in the Pasteur Institute study [22], we excluded from this analysis the dynamics of the COVID-19 epidemic within the retirement homes for aged people in France, as both the demographics of this part of the population and the transmission dynamics are very specific, while the data used to describe the COVID-19 dynamics in this setting is very uncertain.

The individuals having arrived in the *R_S_* or *R_M_* classes then have a period of immunity of 1/γ*_R_* days and then can start a new infection again either in the class *S_f_* (”Susceptibles Fragilized”) or in the class *S_r_* of susceptibles (having had a first infection) with no particular fragility. The probability to arrive in class *S_f_* is taken to be *f*. Each of the classes *S_f_* and *S_r_* is then the starting point of a new infection pathway of the same type as before. In all notations the suffix “i” in a class name will denote the “initial” infection block, the suffix “f” the “fragilized” pathway and the suffix “r” will denote the ’reinfection” branch.

Mathematically the SEIRIS model is written for *a* ∈ *{i,f,r}* (see figures 2 and 3 for illustrations):

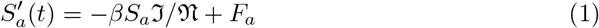

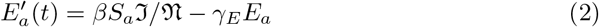

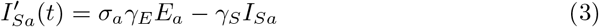

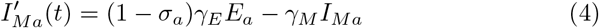

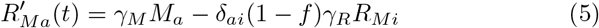

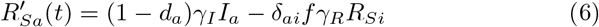

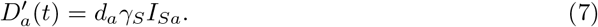

with the notations

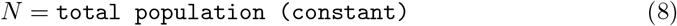

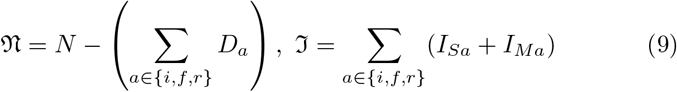

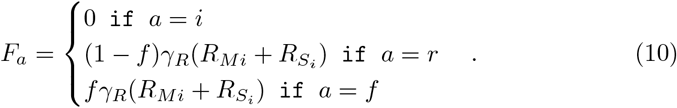

Note that the last term in equations (5) and (6) is only non-null for the class ’i’ and represents the outgoing flow of primary infected individuals to the “fragile” or “non-fragile” blocks, see also figure 3. The incoming part of these flows, denoted *F_a_* appear in equation (1).

To model the impact of previous infections on the next possible epidemic we have to take several facts into account: while the SARS 2003 had a negligible asymptomatic class (see [15]) the COVID-19 has non-negligible number of mild or even asymptomatic individuals. Those patients are usually not detected and their estimation is statistically challenging. Even more difficult is to extrapolate the state of the immune system after a mild or severe infection and how this can be translated to future immune unbalanced response. Our assumption is the following: a coronavirus infection will sensitize a proportion of *f* % infected persons to future infections; for these persons, the immune function parameters *σ* and *d* will increase.

### 2.2 Choice of parameters

The parameters of the model were set as follows: the scenario independent infection parameters were fit, once for all, to match the curve of cumulative fatalities in a model without reinfection and fragilization (i.e., only the first block present). The results are given in tables 1,2 and figure 1. Note that *R_t_* is only known up to time when data is available (that was stopped at August 18th). Concerning future values of *R_t_* we took the following assumption: the *R_t_* value will be set constant, at mid-distance between epidemic extinction (*R_t_* < 1.0) and the government threshold for drastic interventions *R_t_* = 1.5), i.e. *R_t_* = 1.25. Once the initial simulation has been calibrated, we moved to the challenge. The set of scenarios considered is given in table 3.

**Table 1:** Description and values of the parameters used in the simulations.

| parameter | description (unit) | value (95% CI if available) |
| --- | --- | --- |
| $\beta$ | transmission rate | from formula (12) |
| $\sigma_i$ | severe infection rate | 3.6% (2.1%–5.6%) [22] |
| $\gamma_E$ | incubation time (day<sup>-1</sup>) | 0.25 (fit to data) |
| $\gamma_M$ | infectious time, mild infections (day<sup>-1</sup>) | 0.17 (fit to data) |
| $\gamma_S$ | infectious time, severe infections (day<sup>-1</sup>) | 0.14 (fit to data) |
| $d_i$ | Fatality rate (severe infections) | 18.1% (17.8%–18.4%) [22] |
| $t_0$ | Initial simulation time | Jan 28th 2020 |
| $(I_{Sa}(0), I_{Ma}(0))$ | infected at initial time: (severe, mild) | (1, 37) (fit to data) |
| $\gamma_R$ | immunity time (day<sup>-1</sup>) | 1/142 [26], see also [14] |
| $f, \sigma_f, \sigma_r$ | parameters for | scenario dependent |
| $d_f, d_r$ | other branches | cf. table 3 |
| $R_t$ | effective reproduction rate | fit to data<br>see table 2, figure 1 |

**Table 2:** Values of the effective transmission rates used in the linear interpolation that defines *R_t_*.

|  |  |  |  |  |  |
| --- | --- | --- | --- | --- | --- |
| Date | 1/1/20 | 25/2/20 | 17/3/20 | 31/3/20 | 14/4/20 |
| $R_t$ | 1.7 | 2.9 | 5.49 | 0.68 | 0.61 |
| Date | 28/4/20 | 12/5/20 | 26/5/20 | 9/6/20 | 23/6/20 |
| $R_t$ | 0.53 | 0.64 | 0.66 | 0.59 | 0.76 |
| Date | 7/7/20 | 21/7/20 | 4/8/20 | 18/8/20 | 27/1/21 |
| $R_t$ | 0.84 | 0.82 | 1.25 | 1.25 | 1.25 |

**Table 3:** Scenarios definition (parameters) and results. Baseline scenario has ID=0. All results can be reproduced using the Python code provided as supplementary material.

| ID | fragile<br>($f$) | $(\sigma_f, d_f)$<br>$(\sigma_r, d_r)$ | severe cases<br>from<br>18/08/20 | deaths<br>from<br>18/08/20 |
| --- | --- | --- | --- | --- |
| 0 | 0% | $\sigma_r = \sigma_i, d_f = d_i$ | 150265 | 26863 |
| 1 | 33% | $\sigma_r = 0, \sigma_f = 100\%, d_r = d_f = d_i$ | 200189 | 35770 (+33%) |
| 2 | 33% | $\sigma_r = \sigma_i, \sigma_f = 100\%, d_r = d_f = d_i$ | 204190 | 36483 (+36%) |
| 3 | 100% | $\sigma_f = 100\%, d_r = 10\%$ | 309408 | 41792 (+56%) |

## 3 Results

We compared the outcome, in terms of deaths, of the impact of first infection on the secondary infection (challenge) outcomes under the Cuban hypothesis described in section 1 through several scenarios. The baseline scenario (ID=0) has no fragilization neither long term immunization, immunity from initial infection being lost after 1/γ*_R_* days.

The first alternative scenario is when all people at high risk (age above 65 years or with co-morbidities, i.e. 33% of the population) will experience, upon reinfection, the severe version of the disease; on the contrary all other individuals, upon reinfection, will only experience mild forms. In this scenario the second wave (defined as cases occuring after August 18th) will witness a +33% increase in the death toll. The increase is due to the fragile branch as can be checked considering a second alternative scenario where the non-fragile branch is allowed to experience severe forms (with same rate as the primary infection, i.e., neither fragilization nor long term immunization). In this second scenario the death toll is 36% larger than the baseline, which is not very far from the figure previously obtained.

**Figure 1:**
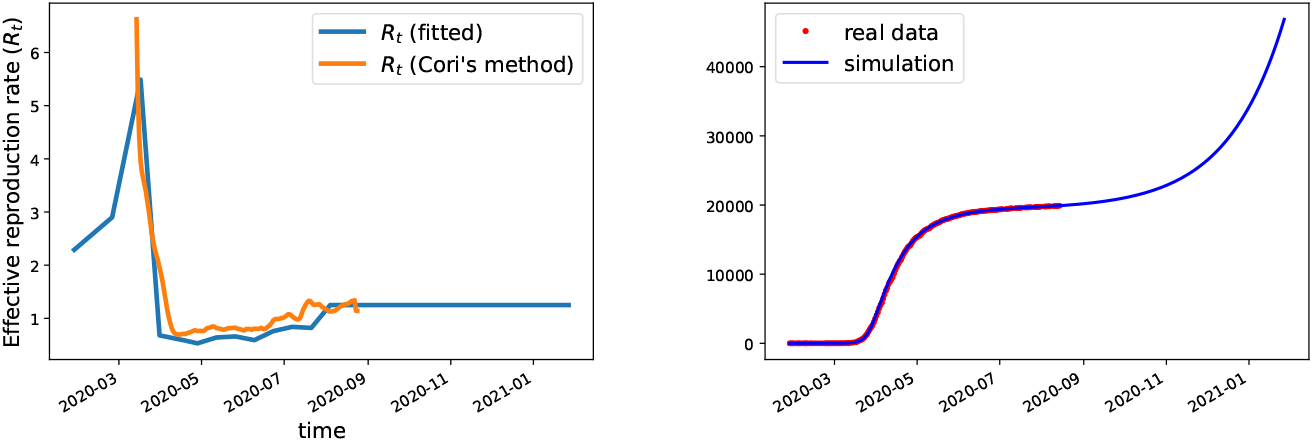
**Left:** Effective reproduction rate *R_t_* used in the simulations compared with the effective transmission rate published by the government site “Santé France” from [1]; this effective reproduction rate is computed using the Cori’s method [25] (averaged between tests and admissions to emergency units). Our effective reproduction rate *R_t_* is computed using a linear interpolation between the values given in table 2 obtained after fitting the cumulative deaths curve until August 18th. **Right:** fit quality (cummulative deaths) produced by this choice of R_t_ (other parameters are as in table 1).

**Figure 2:**
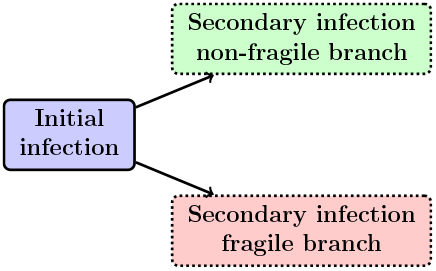
Schematic illustration of the SEIRIS model in equations (1)-(7). See figure 3 for details.

**Figure 3:**
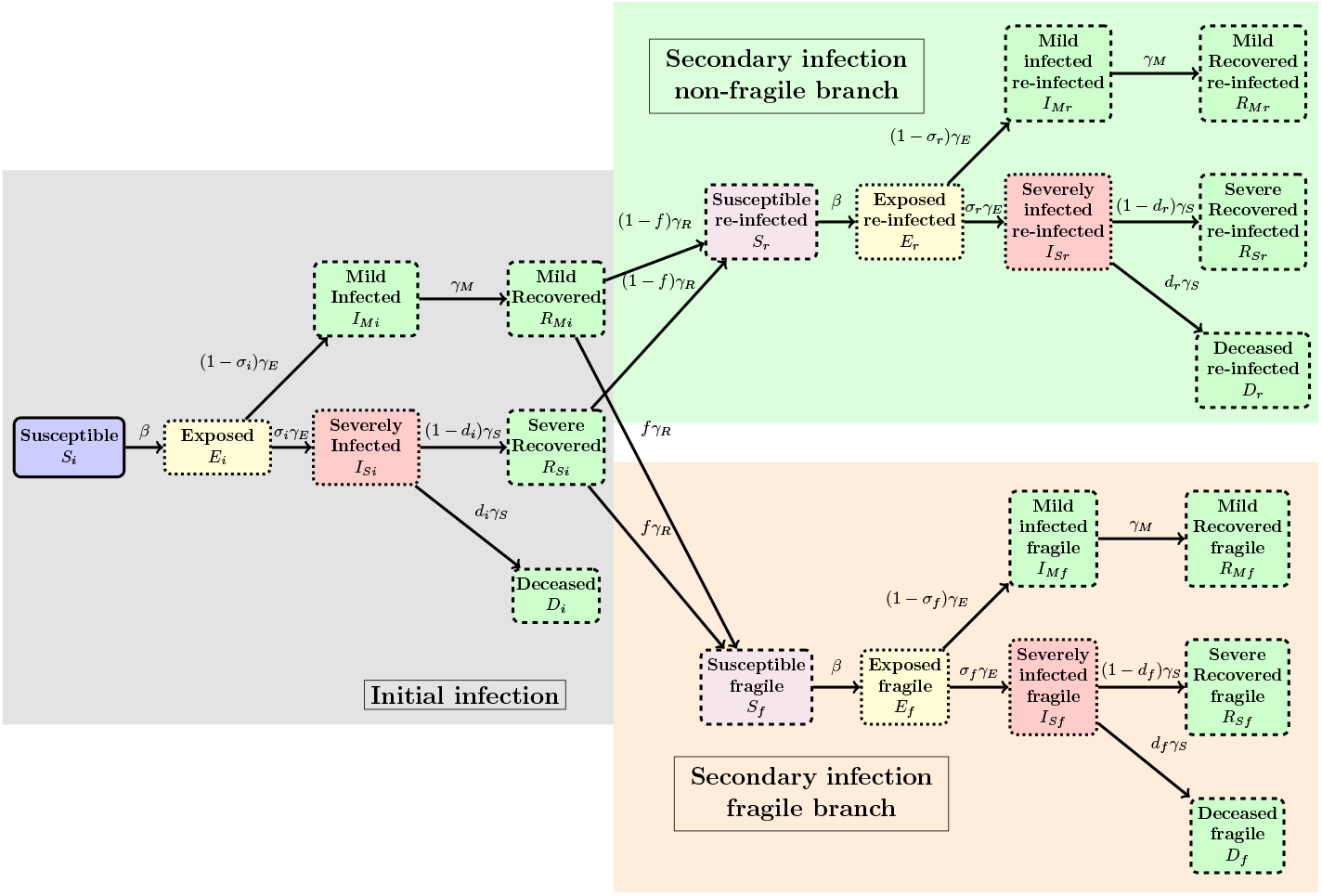
Detailed illustration of the flow of the SEIRIS model in equations (1)-(7). See figure 2 for a brief illustration of the general flow.

The increase between the baseline and the first alternative scenario is due to the reinfection phase: in the baseline scenario, 4.1% of the all deaths are due to the reinfection while in the first alternative scenario, 28% of all deaths come from re-infections, thus the reinfection plays a significant role in augmenting the epidemic burden.

Until now, the scenarios only considered the probability to have a severe form. We investigated next an even more extreme scenario whose parameters are similar to the SARS 2003 epidemic: upon reinfection all individuals experience severe forms with a death rate of 10%. The results show, as expected, a further deterioration of the outcomes with respect to all other scenarios (56% more when compared to the baseline). All results can be reproduced using the Python code provided as supplementary material.

## 4 Discussion and conclusion

We propose an epidemic model to investigate the immune function alteration induced by an initial SARS-CoV-2 infection, similar to the antibody-dependent enhancement phenomenon. Note first that the relatively controlled nature of the 2003 SARS epidemic did not allow us to draw conclusions on how the 2003 epidemic influenced the infected (too few cases); by contrast, if a sensitizing process in the immune response triggered by SARS-CoV-2 exists, the pandemic nature of the 2019/20 COVID-19 outbreak will likely have noticeable effects on the overall population health state. In particular, this implies that additional care has to be observed when validating vaccines against the COVID-19.

We developed a scenario meant to explore the consequences that a previous infection changed the likeliness to get a severe form when challenged by a new infection, possibly also increasing the death risk of severe forms (similar to more documented ADE situations as for dengue, see [12]). Irrespective of the precise parameter values we expect that this could be used as a promising methodology to quantify the epidemiological impact of ADE, with coronaviruses as key examples.

The deterioration of the immune function can be evident on two occasions: either when a second epidemic occurs or when submitted to a challenge. The challenge can be very diverse, not necessarily in the form of a fully-fledged epidemic but also a small infection with an endemic coronavirus. This may manifest in the form of multi-organ inflammation as witnessed in UK, France, Canada and the US with some presentations recalling the Kawasaki syndrome [20, 28, 29].

Until now, the global pattern of lethality of the infection appears to be more or less the same worldwide. To monitor whether an ADE-like scenario is emerging it is critical to identify, very early on, regions in the world where a significant number of young people is affected by a severe form of the disease. If this happens analysis of the viral genomes involved in these cases might reveal key features that trigger ADE.

We include a comparison with the 2003 SARS coronavirus as a mean to illustrate the shifting nature of the age classes of individuals most affected by the virus. Although this is not an evidence per se, it may allow investigators to attract research interest and more quantitative qualification of the dynamics of cross-immunity (or fragility) between SARS-CoV-2 and other (corona)viruses.

We present simulation data that measure the possible impact of such a immune function evolution. We emphasize that the alternatives considered here and the resulting figures should not be viewed as quantitative predictions but as “worst case scenarios” that may motivate further, more clinically oriented, research. Of course, our model has many limitations and the real-life immune dynamics may show a more nuanced behavior in which the immunity could be lost after a certain period of time, renewed under certain levels of new exposures to the virus, or even lost again if the additional exposures are too (or too less) frequent; further investigation into the determinants of the immune dynamics is necessary to unveil the conditions of the correct and impaired immune response. Until this is carried over, past COVID-19 patients should not be considered as having a life-time permanent acquired immunity to SARS-CoV-2 or other coronaviruses.

## Data Availability

all data used is available from public sources that have been cited.

## Acknowledgements

We thank the CNRS MODCOV19 research platform for their support and two anonymous referees for their suggestions.

## A The Cuban hypothesis: Comparing case fatality rates in SARS 2003 and 2019-2020

In the absence of any relevant serological information, we compared age-dependent case fatality ratios of the SARS 2003 and COVID-19. More precisely, using data from [16] (see [6, 11] for additional data) we computed the age-dependent death rate for all 1755 SARS 2003 patients in Hong Kong. The benefit of using this database is that it contains information on all patients and therefore has no representation bias. For COVID-19 such data is not yet available so we used the Institut National d’Etudes Démographiques (INED) database [18] at the date of May 4th 2020. The two bases do not allocate cases exactly to the same age groups, we grouped together the 5-year 2003 data to fit the 2020 INED 10-year database ranges and on the other hand use uniform attribution for ages in the “above 75” 2003 HK class) to fill two corresponding classes “70-79” and “80+”.

The comparison of the 2003 and 2020 data in figure 4 shows a striking difference between a panel of several European countries (France, Spain, Italy) with respect to the Hong Kong results. All curves share a common shape: a constant null range followed by a marked (exponential) increase starting in the “30-39” age group for the 2003 data and “50-59” group for all countries in 2020. This 20 years shift at the onset of the exponential regime can be interpreted as resulting from the existence of a common cause (situated approximately 30-40 years before 2003 and 50-59 years before 2020, that is, between 1960 and 1970) that affected all people living at that date and which could explain why there are, 30 (respectively 50) years later, affected by the coronavirus outbreaks in a more severe way. Since the cause occurred before 1970 people born after this date will have no fragility, which is precisely what is seen in the plot. We interpret this as an evidence that previous infection history can negatively impact the individual outcome of a future infection. Note that historically the 1960-70 decade is rich in epidemic / virus related events: the Hong Kong flu of 1968-1969 (that killed an estimated 1 million people worldwide) but also the start of the identification of the four endemic human coronaviruses (mid-1960s). Note that such a conclusion is consistent with recent investigations, see [8] that showed that individuals are prone to repeated infection with coronaviruses, with the unfortunate consequence that not only the immunity may vanish (sometimes within a year) but that the re-infection can be more severe. Data in figure 4 substantiates the same conclusion.

**Figure 4:**
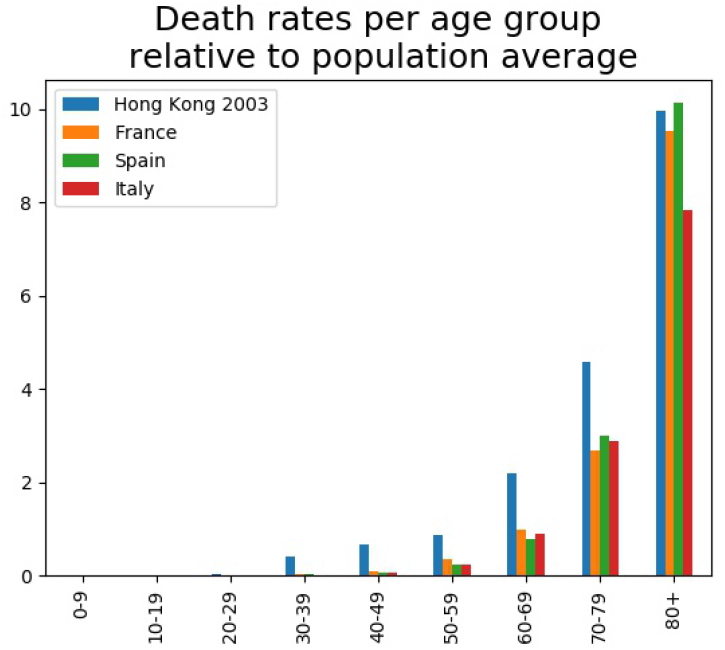
Age dependent death rate comparisons between SARS 2003 and COVID-19.

## B Technical details

We compute here the basic and effective reproduction ratios (for instance the famous “*R*_0_”) of the model in equations (1)-(7).

To this end we recall some facts concerning this computation. We will use the “next generation matrix” method as in [27, 21]. The method uses only the infectious compartments (here x = (*E_i_, I_Mi_, I_Si_, E_f_, I_Mf_, I_Sf_, E_r_, I_Mr_, I*_Sr_) and works with the linearized equation in the form

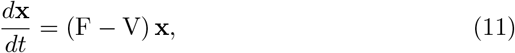

where the matrix F describes the rate of new infections and V describes the transfer between compartments. Then the *basic reproduction ratio* is defined as the largest eigenvalue of FV^−1^. Note that if (λ, *υ*) is an eigenvalue-eigenvector pair for FV^−1^ i.e., FV^−1^ *υ* = λ*υ*, then F*w* = λV*w* for *w* = V^−1^*υ*, thus *λ* if a root of the following equation det(F − *λ*V) = 0. The same method works for the effective reproduction ratio.

### Proposition B.1.

*The effective R_t_ reproduction ratio of the model* (1)-(7) *is given by the formula:*

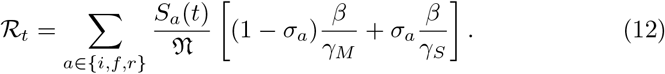

*In particular the basic reproduction ratio R*_0_ *is*

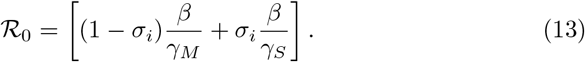

#### Proof.

To obtain (12) we note that, with the previous notations

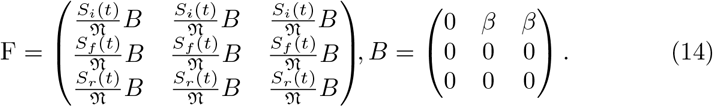

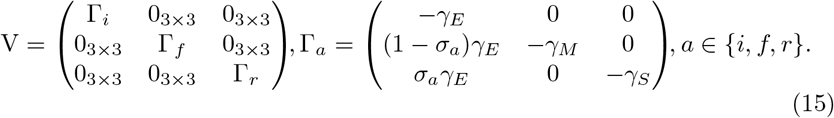

Manipulating the determinant of F− *λ*V we obtain (a symbolic Python code reproducing this computation is provided as supplementary material):

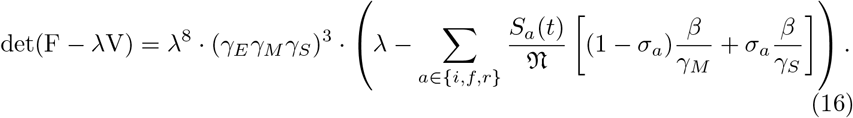

Thus the equation det(F − λV) = 0 has 8 null roots and the only non-null root is real and given by the formula (12).

The derivation of (13) from (12) is immediate from the fact that 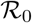 corresponds to a fully susceptible population i.e., 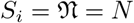 and *S_f_* = *S_r_* = 0.

## Conflict of interest

The authors declare no conflict of interest.

## Notes

### Competing Interest Statement

The authors have declared no competing interest.

### Funding Statement

Logistic support from the MODCOV19 platform is acknowledged.

### Author Declarations

Research only involves theoretical epidemiological data, no IRB oversight is required.

### Summary of Updates

Model explicitly shown in illustration, several scenarios added.

